# The Inflation Reduction Act’s Impact Upon Late-Stage R&D

**DOI:** 10.64898/2026.05.20.26353648

**Authors:** Harry P. Bowen, Gwen O’Loughlin, Claire Schleicher, Duane G. Schulthess

## Abstract

**Background:** The impact of the Inflation Reduction Act (IRA) upon late-stage developments has been assumed to be limited. The Congressional Budget Office’s IRA analysis excluded post-approval innovation, potentially overlooking substantial economic risks to drug developers and declines in the availability of treatments in areas of high unmet medical need such as oncology.

**Methods:** A total of 1148 secondary trials from 364 FDA-approved medicines, published from 2018 to 2025, were obtained from Biomedtracker and clinicaltrials.gov. Using fractional multinomial logit, we model the share distribution of secondary indication studies across 19 disease groups and assess the change in this distribution post-IRA. We also assessed the number of secondary treatment studies pre- vs. post-IRA using multiple linear regression.

**Results:** After the IRA’s introduction, small molecule follow-on studies in oncology exhibited a statistically significant 35% decline (R^2^ = .48, p < 0.014) and lead indication small molecule oncology approvals exhibited a statistically significant 27% decline (R^2^ = .70, p < 0.002). We also find a statistically significant 14% decline in the share of orphan oncology studies pre- vs. post-IRA (p<0.001).

**Research Conclusions:** This study’s results refute claims that the IRA would have minimal negative effects on patient access or late-stage biopharmaceutical R&D. We hope this study reinvigorates debate about the law’s unintended consequences and encourages thoughtful policy solutions, as the IRA manifestly creates disincentives that negatively impact patients seeking needed new medicines, particularly those requiring cures addressing metastatic late-stage cancers.

## Introduction

Previous research has shown a direct and material reduction in early-stage venture capital and angel investments into small molecules that target indications with high prevalence in the Medicare-aged population after introduction of the Inflation Reduction Act (IRA) on September 27, 2021^1^ . In contrast, IRA’s impact upon late-stage developments has largely been ignored or assumed to be limited. For example, the UK’s Office of Health Economics (OHE) noted the Congressional Budget Office (CBO) excluded late-stage post-FDA approval research from its IRA analysis^2^, stating that the CBO “Relies on a narrow conception of biopharmaceutical innovation that continues to exclude post-approval innovation.”^3^ Similarly, some academics have stated categorically that, “capital already committed will be deployed even if return expectations have shifted.”^4^ Vogal, et al. present evidence that “Late-stage private company and public equity valuations, IPOs, follow-on offerings, and mergers and acquisitions in biopharma largely held steady in the 18 months since passage of the IRA.”^5^

If incorrect, the belief that the IRA had limited impact upon late-stage and follow-on research would ignore potentially substantial economic risks to drug developers as well as potential declines in the availability of treatments in areas of high unmet medical need, such as oncology.

After a drug is approved, it is common for the drugmaker to commit to follow-on research for secondary indications. For example, the PD-1 inhibitor pembrolizumab (Keytruda), initially approved for advanced melanoma in September 2014, is now approved for more than 40 indications for both subcutaneous and intravenous treatments^6^. In prior research^7^, we showed that follow-on indications help ensure a positive net present value (NPV) for the drug developer, thereby allowing the therapy to remain in the market to treat more patients impacted by maladies with unmet medical needs.

Limited research has been conducted on the potential impacts of the IRA upon late-stage and post approval research into follow-on indications. The potential risks to post-approval studies created by the IRA were highlighted by Patterson et al., stating, “Given the role of multiple indications in expanding treatment options in patients with cancer, IRA-related changes to development incentives are especially relevant in oncology.” ^8^They further outline that these IRA risks are particularly acute in oncology due to its heterogeneous clinical development pathways^9^.

This study addresses this gap in the literature by conducting a multi-year, multi-indication analysis of post-approval R&D into secondary indications before and after IRA’s introduction to detect whether the IRA has had measurable impacts in this regard.

## Materials and Methods

To analyze the impact of the IRA legislation on a firm’s choice to develop secondary indications, we classified 1148 secondary trials drawn from 364 approved lead indications of new molecular entities and biological products across 175 firms for the years 2018-2025. Clinical trial data were obtained from Biomedtracker, a subscription-based competitive intelligence platform that tracks the commercial, clinical, and regulatory status of developmental drugs and therapies, and clinicaltrials.gov.

The year of a given secondary indication study was assigned to the year of the lead indication’s FDA approval. As we have annualized data, we chose January 1, 2022, as the date of the IRA’s introduction.

To detect any change in the number of follow-on studies launched after the IRA’s introduction, our cohort of 1148 follow-on studies were assigned to one of nineteen disease groups: Allergy, Autoimmune/Immunology, Cardiovascular, Dermatology, Endocrine, Gastroenterology (Non-IBD), Hematology, Infectious Disease, Metabolic, Neurology, Obstetrics/Gynecology, Oncology, Ophthalmology, Orthopedics, Psychiatry, Renal, Respiratory, Rheumatology (Non-Autoimmune), and Urology.

For each disease group, the probability of a change in the number of secondary treatments studied pre- and post-IRA by its indication, modality (small vs. large molecule), rare disease status, and exposure to the Medicare-aged population was measured using multiple linear regression.

To assess changes in the disease groups targeted pre- and post-IRA, we modeled the probability that a secondary indication study targets a given disease group using a fractional multinomial logit regression (FMLogit) model. The form of the model is:

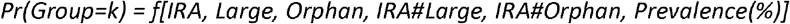

where:

Pr(Group=k) = probability that a study is conducted for disease group “k”.

IRA = 1 if the study commenced post-IRA, IRA = 0 if study began pre-IRA.

Large = 1 if biologic (large molecule), Large = 0 if small molecule.

Orphan = 1 if indication targets rare disease, Orphan = 0 if indication targets non-rare disease.

IRA#Large = interaction between IRA and Large variables.

IRA#Orphan = interaction between IRA and Orphan variables.

Prevalence (%) is the percent of U.S. population aged 65 and older who have the disease targeted by the secondary indication.

The FMLogit model estimates the proportion of secondary studies targeting treatments within a given disease group. For each disease group, the IRA variable indicates whether the probability (proportion) changes following the IRA’s introduction in 2022, and in what direction. The model is akin to the multinomial logit model, but it does not assume that disease group choices are mutually exclusive. By construction, the model ensures that the probabilities (proportions) over all disease groups sum to one.

## Results

We observe a 12% decline in the total number of post-approval studies following the introduction of the IRA, from 610 to 535 studies (Figure 1). This decline in follow-on studies is particularly pronounced in oncology, which evidences a 20% decline (from 325 to 260 studies).

**Figure 1.**
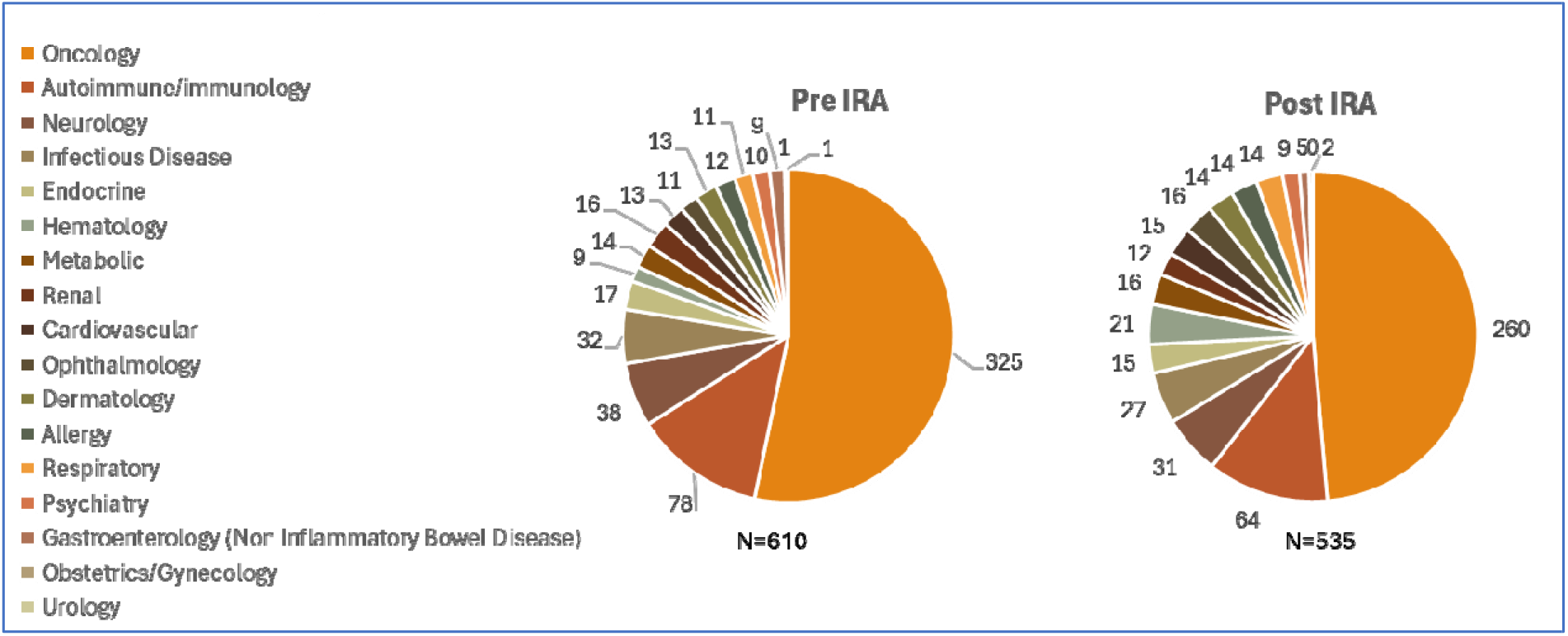
Shares of secondary research studies pre- vs. post IRA, 2018 – 2025, n = 1145. Oncology studies see the largest decline in total after the IRA’s introduction. Sources: Biomedtracker, ClinicalTrials.gov.

While we observe a decline in the total number of follow-on studies, oncology is the primary driver of this reduction, as over 50% of the 1145 secondary studies in our cohort were in oncology. Large declines in the number of active follow-on oncology studies are observed in 2021 & 2022, contributing to a continual downward linear trend through 2025 (Figure 2).

**Figure 2.**
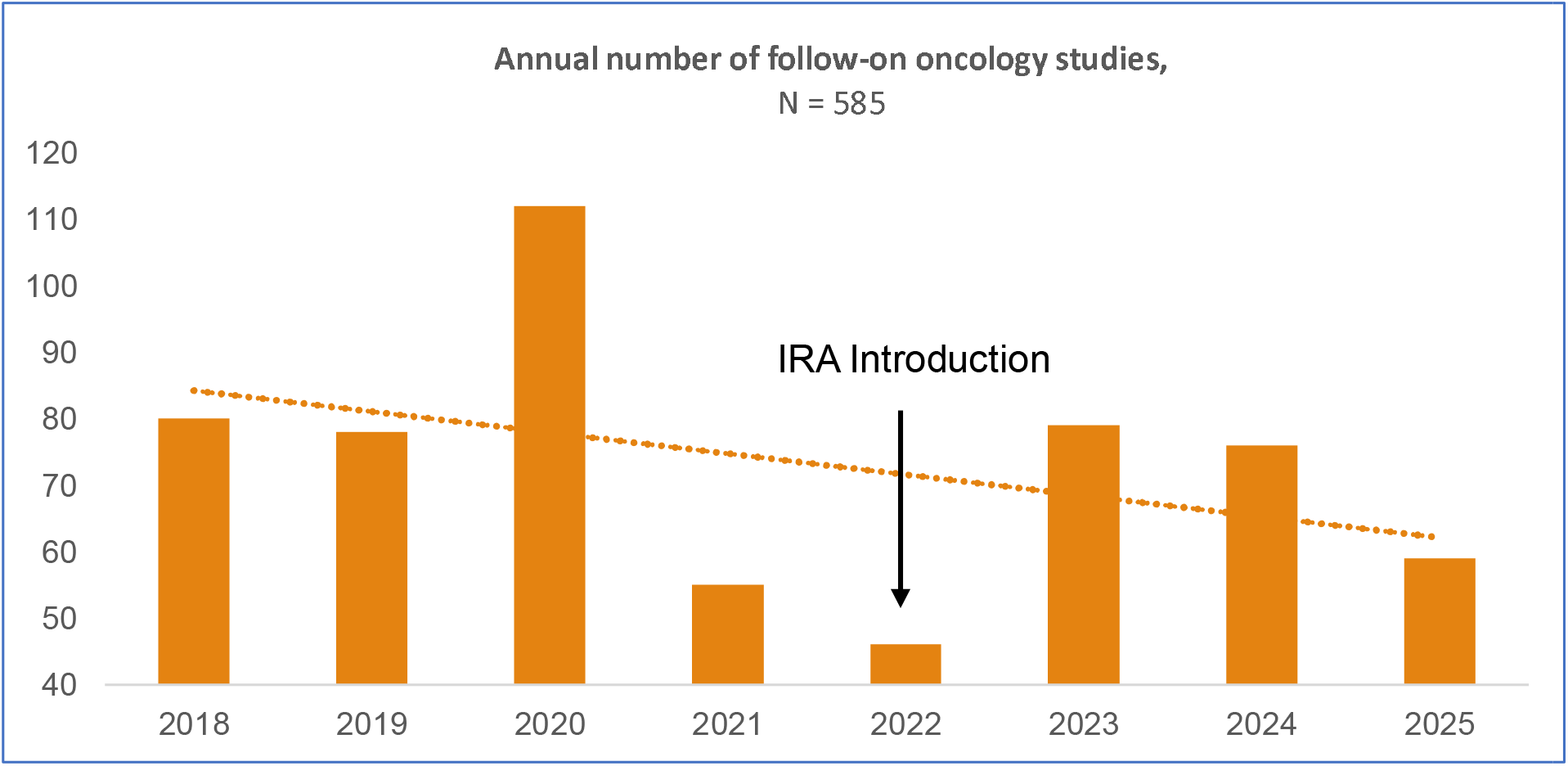
Number of follow-on oncology studies, 2018-2025. Sources: Biomedtracker, ClinicalTrials.gov.

The above analysis does not consider follow-on study modality (small vs. large molecule). Previous studies have shown that the IRA’s differential timelines toward drug price negotiation program eligibility, whereby small molecules are negotiated at 9 years and large molecules at 13 years, disproportionately disincentivizes post-approval research into small molecule drugs.^10,11^

To investigate this issue, we used a multiple regression to predict the impact of the IRA’s introduction on the number of follow-on oncology studies per year when segmented by small vs. large molecules (Figure 3). Small molecule follow-on studies exhibit a statistically significant 35% decline after IRA’s introduction (R^2^ = .48, p<0.014), from 53 launched per year before IRA to 34 afterward. Conversely, we observe no statistically significant change in large molecule (biologic) follow-on studies; the number of these studies is unchanged at 30 per year.

**Figure 3.**
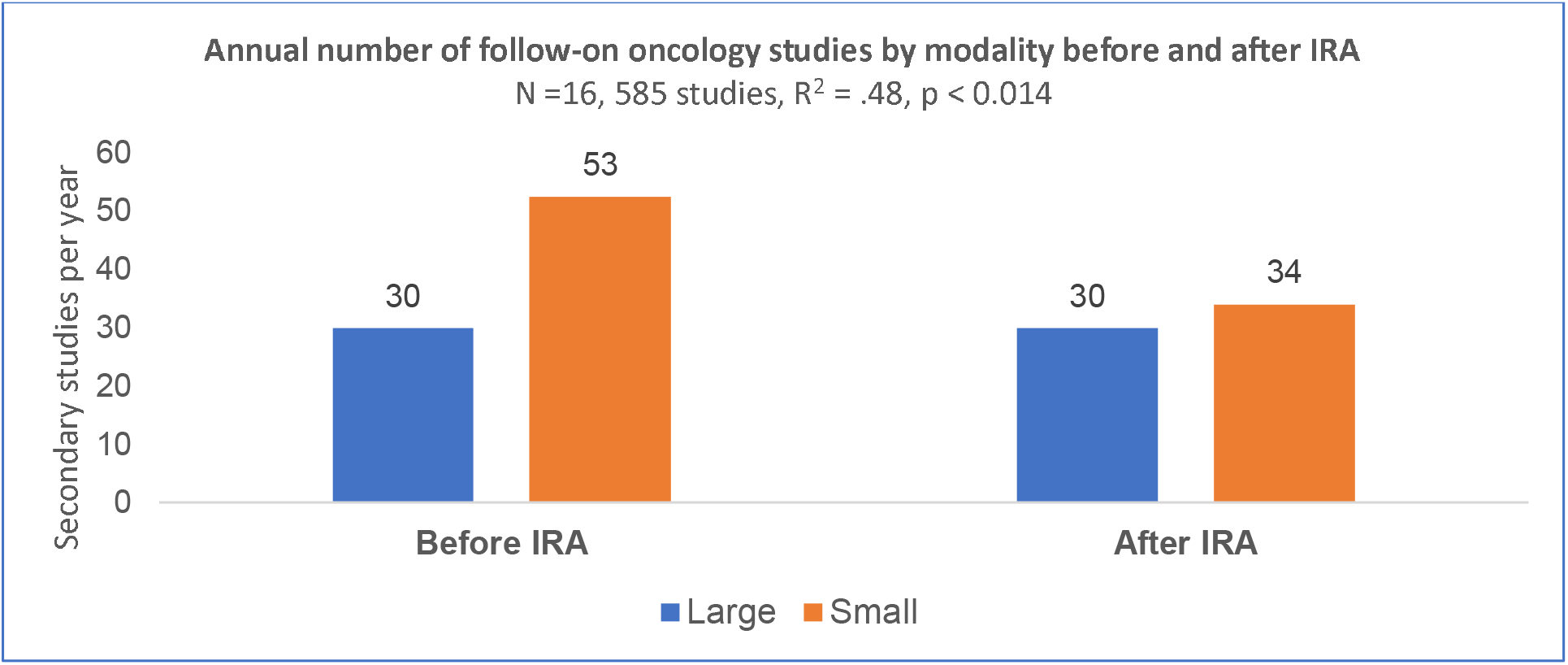
Small molecule follow-on studies exhibit a 35% decline after IRA’s introduction, from 53 per year before IRA to 34 per year after IRA. Large molecule studies are unchanged by IRA. See Statistical Appendix for regression results. Sources: Biomedtracker, ClinicalTrials.gov.

Observing the decline in secondary oncology studies, we used a multiple regression to determine the relationship between the annual approval of lead indication oncology products by modality, before and after IRA’s introduction (Figure 4). Lead indication oncology approvals are the only disease group where we find a statistically significant impact of the IRA after its introduction.

**Figure 4.**
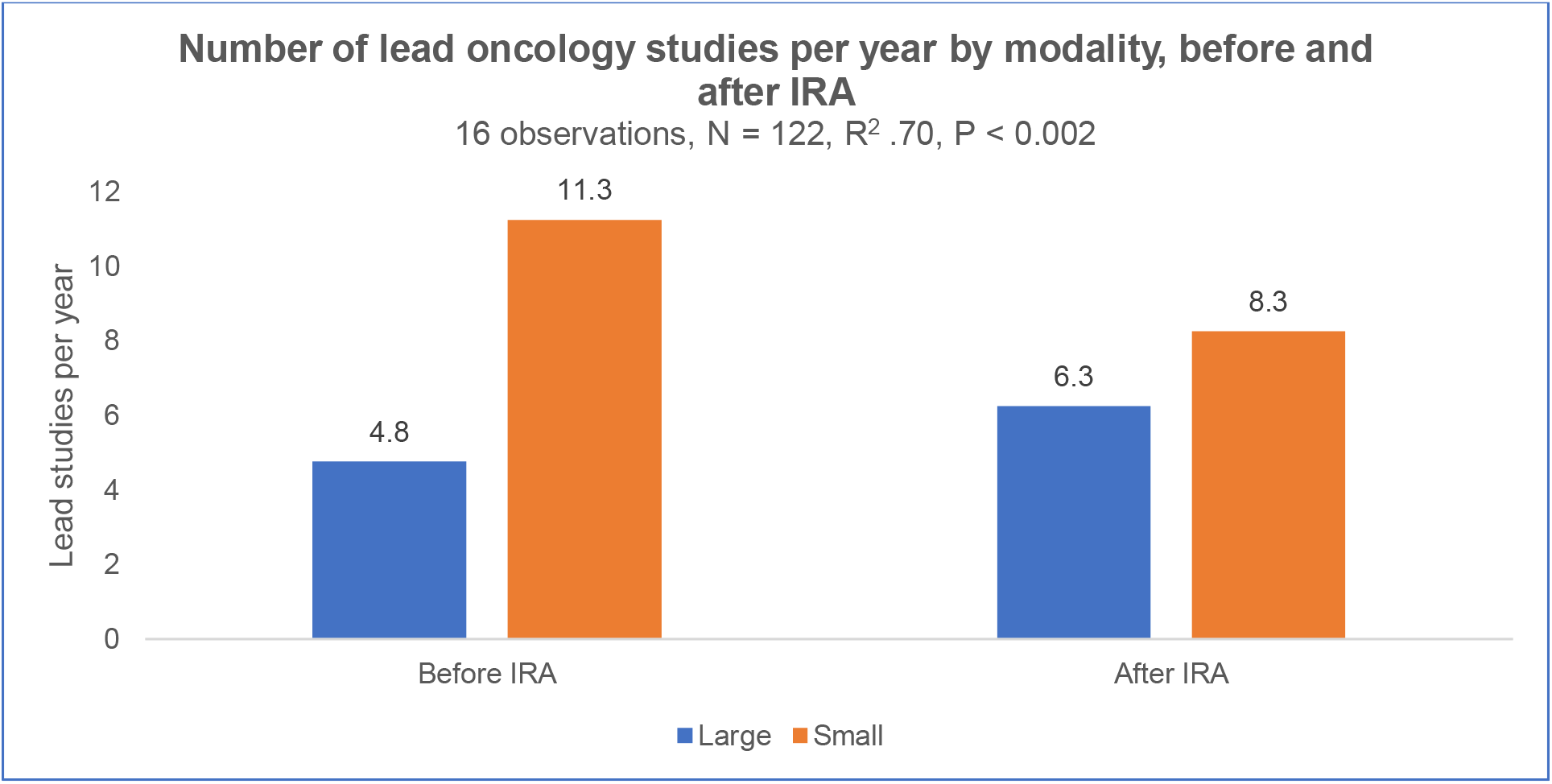
Frequency of lead indication approvals in oncology before and after IRA’s introduction. Small molecule oncology approvals declined by 27% and large molecule oncology approvals increased by 32%. See Statistical Appendix for regression results. Sources: Biomedtracker, ClinicalTrials.gov.

Our regression results indicate that lead indication small molecule oncology approvals declined from 11.3 per year to 8.3 per year after IRA’s introduction, a 27% reduction, and large molecule oncology approvals increased from 4.8 to 6.3 per year after IRA’s introduction, a 32% increase (R^2^ = .70, p < 0.002). As our ratio of small molecule follow-on studies to lead indication approvals in oncology before IRA’s introduction was 4.7:1, we can estimate a four-year decline of 56.3 fewer follow-on studies after IRA’s introduction by the ratio to approved lead small molecule oncology indications.

Our regression results, based on Figure 3, show a decline of 19 follow-on studies per year after IRA’s introduction, representing an aggregate 4-year decline of 76 total studies with a 95% confidence interval of 46 to 226 studies. Our above estimate of a decline in follow-on studies of 56.3, due to the ratio of fewer lead small molecule oncology approvals after the IRA’s introduction, falls within this estimate.

We also conducted the analyses described above for the hematology studies. Specifically, a linear regression predicting the number of secondary hematology studies per year pre- vs. post-IRA was estimated using 12 observations. This indicated a statistically significant post-IRA increase in the number of follow-on hematology studies per year (R^2^ = .39, p < 0.031), which represents an increase of over 200%. However, this increase comes from a low pre-IRA base of 1.5 hematology studies per year.

Finally, a linear regression segmenting hematological follow-on studies by modality (small vs. large molecules) was not statistically significant. Hence, aside from oncology and hematology, no other disease group showed statistically significant changes in the number of follow-on studies per year from pre- to post-IRA.

To examine for pre- vs. post-IRA changes in the “portfolio” mix of studies across disease groups, we estimated the FMLogit model relating the proportion of secondary studies launched in each disease group to the post-IRA variable, and to variables for various characteristics of each therapy (e.g., modality, rare disease status; see Statistical Appendix) .^12^

Comparing pre- vs. post-IRA, we found a statistically significant decline (p<0.001) in the proportion of small molecule oncology studies; a rise in large molecule studies post-IRA was not statistically significant (p<0.416). We also found a statistically significant difference (p<0.0001) in the proportion of large vs. small molecule oncology secondary studies post-IRA, but no significant difference (p< 0.198) between the proportion of large vs. small molecule oncology secondary studies pre-IRA (Figure 5).

**Figure 5.**
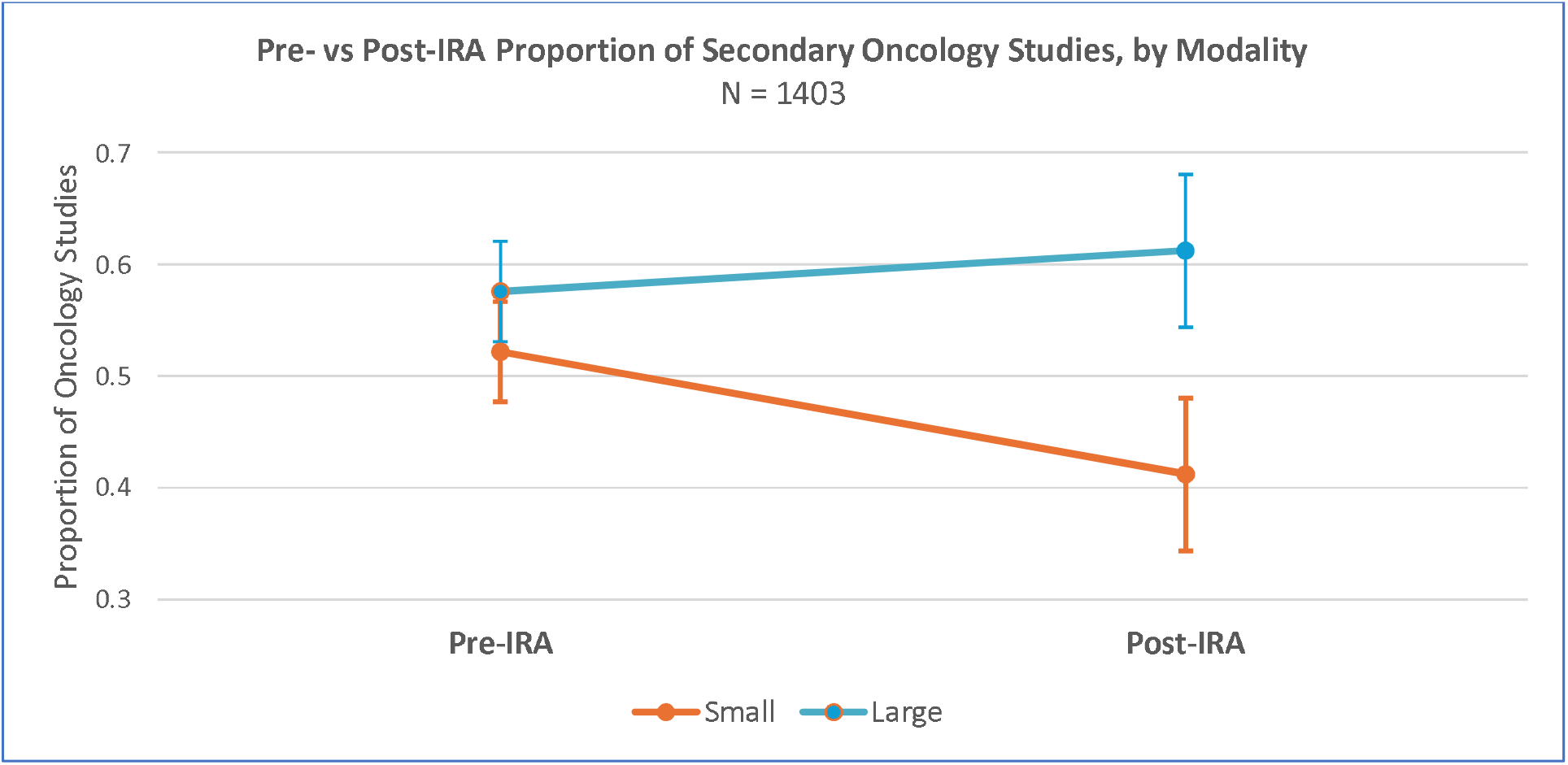
Proportion of secondary oncology studies by modality pre- vs. post-IRA predicted using FMLogit model. Vertical lines are 95% confidence intervals. The pre- vs. post-IRA decline in the proportion of small molecule oncology studies is significant (p<0.001). See Statistical Appendix for contrasts of predicted proportions. Sources: Biomedtracker, ClinicalTrials.gov.

Testing for changes pre- vs. post-IRA in the proportion of oncology follow-on studies by rare disease (orphan) status, we found a statistically significant 14% decline in the proportion of orphan oncology studies post-IRA (p<0.001). In contrast, there was no statistically significant change (p<0.917) in the proportion of non-orphan studies pre-IRA vs. post-IRA (Figure 6).

**Figure 6.**
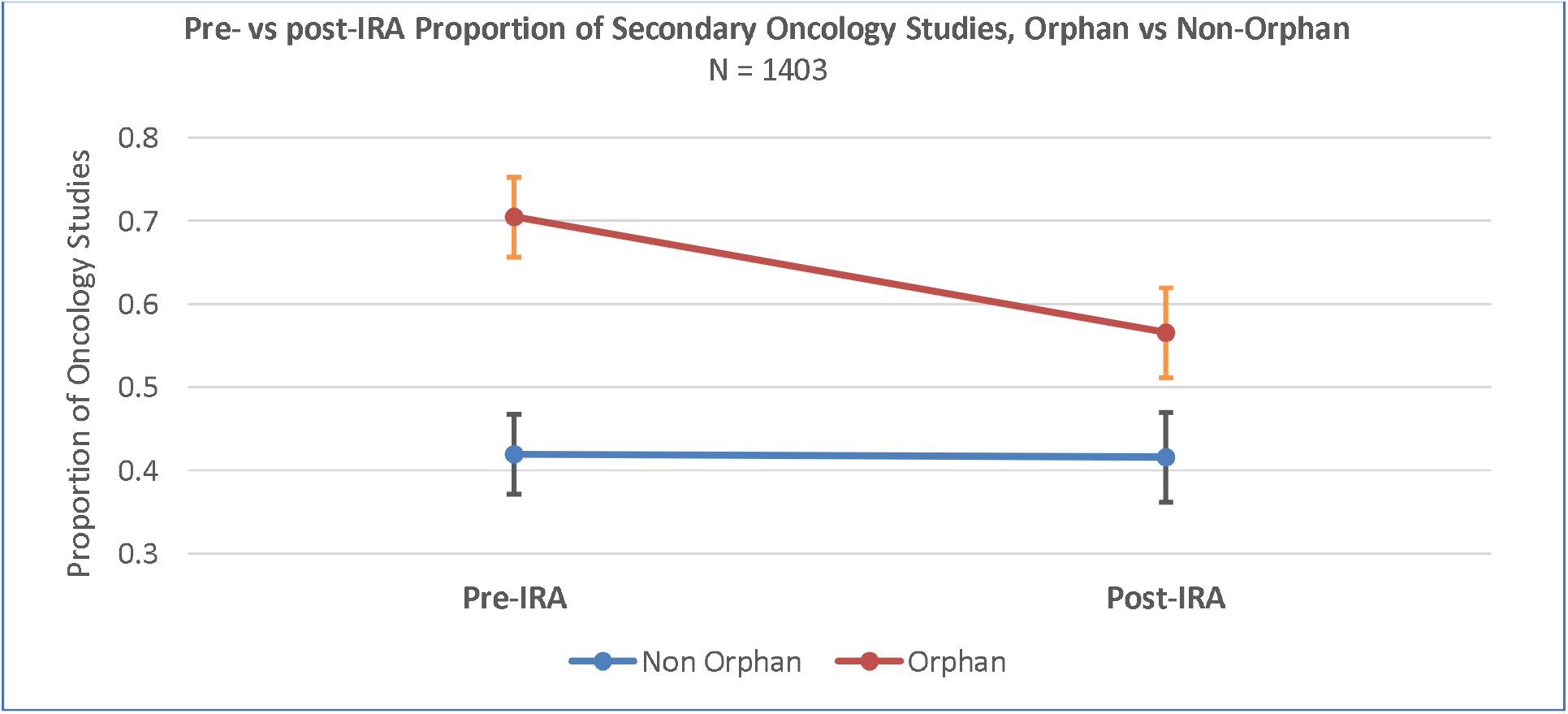
pre-vs. post-IRA predicted proportion of follow-on oncology studies by rare disease (orphan) status using FMLogit model. Decline in follow-on orphan studies is statistically significant (p < 0.001). See Statistical Appendix for contrasts of predicted proportions. Sources: Biomedtracker, ClinicalTrials.gov.

Using the FMLogit model, we also tested for a change in the proportion of secondary hematology studies by modality. We found a statistically significant increase in the proportion of small molecule follow-on hematology studies (p<0.02) post-IRA. We also found a statistically significant difference in the proportion of small vs large hematological secondary studies post-IRA (p<0.0001), and a marginally statistically significant difference pre-IRA (p<0.078).

Testing for an association between orphan status and follow-on hematological studies, we found a statistically significant higher proportion of orphan vs. non-orphan hematology follow-on studies (p<0.009) post-IRA. We also found a marginally significant increase (p<0.051) in the proportion of orphan hematology secondary studies pre- vs. post-IRA. Other than for oncology and hematology, none of the other disease groups evidenced a statistically significant impact of the IRA, whether in total or segmented by modality^13^ .

## Discussion

While late-stage research has been assumed to be resistant to the disincentives of IRA price negotiations, this research has found evidence that this assumption is likely erroneous. We find substantial and material changes to drug discovery post-approval research pipelines after the IRA’s introduction, manifested particularly in significant declines in the late-stage oncology research pipeline.

The primary driver of these declines was found to be in small molecule oncology studies, which comprise a third of all lead indications and over half of all secondary post-FDA approval studies, with the latter declining 35% after the introduction of the IRA. This decline in small molecule oncology studies following IRA’s introduction raises concerns about long-term innovation and patient access to new therapies—particularly for those with advanced or treatment-resistant cancers.

Interestingly, these findings mirror many of those found in our previous publication investigating changes in early-stage VC and angel investments pre- vs. post-IRA^14^. Given this, the assumption that the IRA would not impact late-stage research pipelines may be an example of the sunk-cost fallacy, whereby an investment with a low payoff probability compared to other potential future opportunities is continued because an earlier investment was made^15^. To the contrary, this study provides evidence that late-stage drug development has in fact been substantially altered in response to the risks of negotiation under the IRA.

In addition to the marked reductions in late-stage oncology studies, we also observe a statistically significant increase in the number and proportion of secondary hematology studies per year. However, we find no differentiation by modality, and this increase rises from a low pre-IRA base of just 1.5 hematology studies per year.

Given that our cohort includes data inclusive through December 31, 2025, our findings are unlikely to be substantially influenced by the Orphan Cures Act^16^ passed as part of the Federal Budget reconciliation process on July 4, 2025. Our research finds a significant 14% decline in the proportion of follow-on orphan oncology studies launched after IRA’s introduction. The law excludes orphan drugs from market approval calculations, as well as those that treat more than one rare disease or condition^17^. This provision could likely arrest the decline in small molecule orphan oncology follow-on research studies. However, it is increasingly under pressure to be repealed.

CBO’s initial assessment of the Orphan Cures Act estimated its cost to be $5 billion over10 years. On October 20, 2025, they submitted a revision stating, “The new law will affect price negotiations for several orphan drugs not originally included in the estimates of budgetary effects of section 71203 of the 2025 reconciliation act. After incorporating those drugs into its analysis… the 10-year cost of the section will be $8.8 billion.”^18^ This revision has led to the introduction of legislation to overturn the Orphan Cures Act, which we would expect, if passed, to continue the decline in late-stage orphan cancer treatment research^19^.

### Limitations

As secondary oncology studies are roughly 50% of our cohort, and the mean time to approve a secondary oncology indication is roughly 2 years,^20^ our study design, which ascribes the year of follow-on research to the year a drug’s lead indication is approved, could bias downward the count of secondary studies after the IRA’s introduction. However, our regression analysis of the ratio of lead small molecule oncology approvals to secondary follow-on studies demonstrated that the 27% decline in late-stage small molecule lead indication oncology approvals is directly linked to the observed decline in secondary follow-on studies, a result within our estimated 95% confidence intervals.

Whilst the IRA was initially introduced as the Build Back Better Act on September 27^th^, 2021^21^, we opted in this study to compare four equal years of data before and after the IRA’s introduction. Given the large reductions in R&D activity in 2021, our choice to use January 1, 2022, as our IRA index date may underestimate the impact of the legislation’s introduction.

In contrast to our previous study examining IRA’s impact on early-stage research by a specific indication’s exposure to the Medicare-aged population^22^, this study’s analysis of 389 separate indications yielded no changes in behavior due to a drug’s likelihood to be negotiated under the IRA other than by small or large molecule modality. We interpret this as evidence that companies are broadly pivoting away from small molecule secondary oncology indications, given both their higher likelihood of IRA price negotiation and their exposure to the nine-year pill penalty for small molecules in general when compared to the 13-year negotiation provisions for large molecules^23^.

## Conclusion

The belief that the IRA would have a limited impact on late-stage and post-FDA approval trials appears mistaken; this study presents evidence of significant reductions in follow-on small molecule oncology studies broadly, particularly in late-stage studies targeting orphan cancer indications following the introduction of the IRA. In fact, such impacts were suggested in 2024 by Grabowski and Long, who stated: “Post-approval trials for small molecules, longer-duration trials, and larger-enrollment trials, and post-approval indications focused on limited patient populations and older patients could face particular economic challenges.”^24^

The statistically significant changes found in this study for both lead and secondary oncology studies late in the development pipeline should give regulators pause, particularly those who strongly defended the IRA legislation and asserted that it would have minimal negative effects on patient access or biopharmaceutical R&D. We hope this study reinvigorates debate about the law’s unintended consequences and encourages thoughtful policy solutions, as the IRA manifestly creates disincentives that negatively impact patients seeking treatments in areas of high unmet medical need, particularly those requiring cures addressing metastatic late-stage cancers.

## Supporting information

Statistical Appendix

## Data Availability

Statistical data produced are available online

https://1drv.ms/b/c/fd1ceff1664dae51/IQD7ZO139SuXQ6i_XkPEwdXjAT9sdPaJNm-BzJPlpcKr6Js?e=S4GZIG

## Endnotes

1 Schulthess, D.G., O’Loughlin, G., Askeland, M. et al. The Inflation Reduction Act’s Impact Upon Early-Stage Venture Capital Investments. Ther Innov Regul Sci 59, 769–780 (2025). 10.1007/s43441-025-00773-3

2 https://www.cbo.gov/system/files/2025-04/61231-Drug-Development-Model.pdf (accessed 5/16.26)

3 https://www.ohe.org/insights/is-the-revised-congressional-budget-office-model-of-new-drug-development-fit-for-purpose/ (accessed 5/15/26)

4 “Biopharma Venture Capital And The Inflation Reduction Act,” Health Affairs Forefront, March 5, 2024. DOI: 10.1377/forefront.20240229.164848

5 Vogel M, Conti RM, Chandra A. Biopharma venture capital and the inflation reduction act. Health Aff Forefr. 2024. 10.1377/forefront.20240229.164848.

6 https://www.keytrudahcp.com/approved-indications/ (accessed 5/15/26)

7 https://vitaltransformation.com/wp-content/uploads/2022/11/AA-Project_FINAL_2022_11_11.pdf xPage 21, (accessed 5/15/26)

8 Patterson, J.A., Motyka, J., Salih, R. et al. Subsequent Indications in Oncology Drugs: Pathways, Timelines, and the Inflation Reduction Act. Ther Innov Regul Sci 59, 102–111 (2025). 10.1007/s43441-024-00706-6

9 Ibid.

10 Schulthess, D.G., O’Loughlin, G., Askeland, M. et al. The Inflation Reduction Act’s Impact Upon Early-Stage Venture Capital Investments. Ther Innov Regul Sci 59, 769–780 (2025). 10.1007/s43441-025-00773-3

11 Hanke Zheng, Julie A Patterson, Jonathan D Campbell, Early impact of the Inflation Reduction Act on small molecule vs biologic post-approval oncology trials, Health Affairs Scholar, Volume 3, Issue 8, August 2025, qxaf152, 10.1093/haschl/qxaf152

12 For this analysis, the FMLogit model was estimated using three disease groups: oncology, hematology, and all other groups combined. This aggregation was done after estimating the model for 15 disease groups plus one “other” group that combined the remaining four groups and observing that only the oncology and hematology group equations showed overall significance. See Statistical Appendix for estimation results.

13 The increases in the proportion of hematology studies is not unexpected since the proportions across disease groups sum to one. Hence, a decline in the proportion of oncology studies would be expected to be accompanied by an increase in the proportion of non-oncology studies.

14 Schulthess, D.G., O’Loughlin, G., Askeland, M. et al. The Inflation Reduction Act’s Impact Upon Early-Stage Venture Capital Investments. Ther Innov Regul Sci 59, 769–780 (2025). 10.1007/s43441-025-00773-3.

15 Tait V, Miller HL Jr. Loss Aversion as a Potential Factor in the Sunk-Cost Fallacy. Int J Psychol Res (Medellin). 2019 Jul-Dec;12(2):8–16. doi: 10.21500/20112084.3951. PMID: 32612790; PMCID: PMC7318389.

16 https://www.congress.gov/bill/119th-congress/house-bill/946/all-info (accessed 5/16/26)

17 Ibid.

18 Revised Estimate of Changes Under the 2025 Reconciliation Act for Exemptions From Medicare Price Negotiations for Orphan Drugs, October 20, 2025, Cost Estimate CBO responds to questions regarding the orphan drug exemption under Public Law 119-25, the 2025 reconciliation act. https://www.cbo.gov/publication/61818 (accessed 5/16/26)

19 https://www.forbes.com/sites/joshuacohen/2025/11/04/dems-seek-to-repeal-orphan-only-drugs-exemption-from-medicare-price-negotiations/ (accessed 5/16/26)

20 Patterson, J.A., Motyka, J., Salih, R. et al. Subsequent Indications in Oncology Drugs: Pathways, Timelines, and the Inflation Reduction Act. Ther Innov Regul Sci 59, 102–111 (2025). 10.1007/s43441-024-00706-6

21 https://www.congress.gov/bill/117th-congress/house-bill/5376/summary/00 (accessed 5/19/26)

22 Schulthess, D.G., O’Loughlin, G., Askeland, M. et al. The Inflation Reduction Act’s Impact Upon Early-Stage Venture Capital Investments. Ther Innov Regul Sci 59, 769–780 (2025). 10.1007/s43441-025-00773-3

23 Philipson, T.J. et al., The Potentially Larger Than Predicted Impact of the IRA on Small Molecule R&D and Patient Health, University of Chicago Policy Brief, https://ecchc.economics.uchicago.edu/files/2024/06/Small-Molecule-Paper-20240625.pd (accessed 5/19/26)

24 Grabowski, H., & Long, G. (2024). Post-approval indications and clinical trials for cardiovascular drugs: some implications of the US Inflation Reduction Act. Journal of Medical Economics, 27(1), 463–472. 10.1080/13696998.2024.2323903

