## Supplementary material for "The Inflation Reduction Act’s Impact Upon Late-Stage R&D": Statistical Appendix



### Table A1 – Regression testing impact of IRA on secondary studies for oncology indications by modality

#### Fit

| | |
|---|---|
| N | 16 |
| Mean of Y | 36.6 |

| | |
|---|---|
| Equation | Count of Drug Indication ID = 29.88 + 22.63 Drug Classification Ordinal - 18.5 IRA Interaction |

| | |
|---|---|
| $R^2$ | 0.482 |
| $R^2$ adjusted | 0.402 |
| RMSE | 10.76 |

| Parameter | Estimate | 95% CI | SE | t | p-value | VIF |
|---|---|---|---|---|---|---|
| Constant | 29.88 | 21.65 to 38.10 | 3.8052 | 7.85 | <0.0001 | - |
| Drug Classification Ordinal | 22.63 | 8.386 to 36.86 | 6.5908 | 3.43 | 0.0045 | 1.50 |
| IRA Interaction | -18.50 | -34.94 to -2.059 | 7.6104 | -2.43 | 0.0303 | 1.50 |

| Source | SS | DF | MS | F | p-value |
|---|---|---|---|---|---|
| Difference | 1400.1 | 2 | 700.0 | 6.04 | 0.0139 |
| Error | 1505.9 | 13 | 115.8 | | |


© Vital Transformation, LLC 2026

#### Table A2 – Regression testing IRA's Impact on lead indication oncology studies by modality

##### Fit

| | |
|---|---|
| N | 16 |
| Mean of Y | 7.6 |

| | |
|---|---|
| Equation | Count of Drug Indication ID = 4.75 + 6.5 Drug Classification Ordinal - 4.5 IRA Interaction + 1.5 After IRA |

| | |
|---|---|
| $R^2$ | 0.698 |
| $R^2$ adjusted | 0.622 |
| RMSE | 1.85 |

| Parameter | Estimate | 95% CI | SE | t | p-value | VIF |
|---|---|---|---|---|---|---|
| Constant | 4.750 | 2.736 to 6.764 | 0.92421 | 5.14 | 0.0002 | - |
| Drug Classification Ordinal | 6.500 | 3.652 to 9.348 | 1.3070 | 4.97 | 0.0003 | 2.00 |
| IRA Interaction | -4.500 | -8.527 to -0.4726 | 1.8484 | -2.43 | 0.0315 | 3.00 |
| After IRA | 1.500 | -1.348 to 4.348 | 1.3070 | 1.15 | 0.2735 | 2.00 |

| Source | SS | DF | MS | F | p-value |
|---|---|---|---|---|---|
| Difference | 94.7 | 3 | 31.6 | 9.24 | 0.0019 |
| Error | 41.0 | 12 | 3.4 | | |
| Null model | 135.8 | 15 | 9.0 | | |



© Vital Transformation, LLC 2026

#### Table A3 - Results of estimating FMLogit model predicting log-odds of disease group relative to group “Other”

| | Oncology | | Hematology | |
|---|---|---|---|---|
| **Post-IRA** | -0.236 | [0.218] | 1.431 | [1.013] |
| **Large** | 0.355~ | [0.235] | 1.374+ | [0.786] |
| **IRA#Large** | 0.686* | [0.312] | -1.955+ | [1.013] |
| **Orphan** | 1.672*** | [0.243] | 1.765* | [0.862] |
| **IRA#Orphan** | -0.751* | [0.305] | -0.216 | [1.022] |
| **Prevalence(%)** | 0.055*** | [0.005] | -0.001 | [0.011] |
| **Intercept** | -3.081*** | [0.278] | -4.840*** | [0.941] |
| **Log-Likelihood** | -653 | | | |
| **Chi-Sq.** | 183.2 | | | |
| **Chi –Sq. dof** | 12 | | | |
| **p-value** | 0.000 | | | |
| **N** | 1043 | | | |

~ p<0.15 + p<0.10 * p<0.05 ** p<0.01 *** p<0.001

Notes: Standard errors in brackets. Overall model is statistically significant for explaining the choice of disease group for which a treatment is developed relative to disease group “Other.” Coefficients other than those for interactions indicate the effect of a unit increase in a variable on the log-odds a treatment is developed for the indicated disease group relative disease group “Other”.



© Vital Transformation, LLC 2026

#### Table A4 - Fractional Multinomial Logit, Marginal Effects

| | Oncology | | Hematology | | Other | |
|---|---|---|---|---|---|---|
| **Post-IRA** | -0.062* | [0.026] | 0.018 | [0.009] | 0.043 | [0.027] |
| **Large** | 0.131*** | [0.028] | -0.005 | [0.010] | -0.126*** | [0.028] |
| **Orphan** | 0.212*** | [0.025] | 0.026** | [0.009] | -0.238*** | [0.026] |
| **Prevalence(%)** | 0.010*** | [0.001] | -0.001* | [0.000] | -0.009*** | [0.001] |

~ p<0.15 + p<0.10 * p<0.05 ** p<0.01 *** p<0.001

- For Oncology:
  - The probability of developing an Oncology treatment after the IRA is -0.062 percentage points smaller than it was before the IRA. This effect is statistically significant.
  - The probability of developing a large molecule Oncology treatment is 0.131 percentage points larger than for a small molecule treatment. This effect is statistically significant.
  - The probability an orphan Oncology treatment is developed is 0.212 percentage points larger than for a non-orphan treatment. This effect is statistically significant.
  - A 1 percentage point increase in the prevalence of the targeted indication raises the probability of an Oncology treatment by 0.01 percentage points. Alternatively, a 10-percentage point increase in the prevalence rate would raise the probability of an Oncology treatment by 1 percentage point. This effect is statistically significant.
- For Hematology:
  - The probability of developing a Hematology treatment after IRA is 0.018 larger than it was prior to the IRA. This effect is not statistically significant.
  - The probability of developing a large molecule Hematology treatment is 0.005 6 greater than that of a small molecule treatment. This effect is not statistically significant.
  - The probability of an orphan treatment being developed is 0.026 greater than for a non-orphan treatment. This effect is statistically significant.
  - A 1 percentage point increase in the prevalence of the targeted indication lower the probability of a Hematology treatment by -0.001. Alternatively, a 10-percentage point increase in the prevalence rate lowers the probability of a Hematology treatment by .01 percentage points. This effect is statistically significant.
- One can similarly interpret the effects for Other.


© Vital Transformation, LLC 2026

#### Table A5 - Contrasts of predicted proportion of oncology secondary indication studies by modality

| Contrast | Difference in Predicted Proportions | Std Err | Z-stat | p-value† |
|---|---|---|---|---|
| Pre-IRA Large vs Pre-IRA Small | 0.054 | 0.042 | 1.29 | 0.198 |
| Post-IRA Small vs Pre-IRA Small | -0.110 | 0.033 | -3.33 | 0.001 |
| Post-IRA Large vs Pre-IRA Small | 0.090 | 0.036 | 2.49 | 0.013 |
| Post-IRA Small vs Pre-IRA Large | -0.164 | 0.042 | -3.88 | 0.000 |
| Post-IRA Large vs Pre-IRA Large | 0.036 | 0.045 | 0.81 | 0.416 |
| Post-IRA Large vs Post-IRA Small | 0.200 | 0.037 | 5.46 | 0.000 |

#### Table A6 - Contrasts of predicted proportion of oncology secondary indication studies by rare disease status

| Contrast | Difference in Predicted Proportions | Std Err | Z-stat | p-value† |
|---|---|---|---|---|
| Pre-IRA Orphan vs Pre-IRA Non-Orphan | 0.285 | 0.037 | 7.72 | 0.000 |
| Post-IRA Non-Orphan vs Pre-IRA Non-Orphan | -0.004 | 0.034 | -0.1 | 0.917 |
| Post-IRA Orphan vs Pre-IRA Non Orphan | 0.146 | 0.037 | 3.97 | 0.000 |
| Post-IRA Non-Orphan vs Pre-IRA Orphan | -0.289 | 0.036 | -8.01 | 0.000 |
| Post-IRA Orphan vs Pre-IRA Orphan | -0.139 | 0.038 | -3.65 | 0.000 |
| Post-IRA Orphan vs Post-IRA Non-Orphan | 0.150 | 0.036 | 4.21 | 0.000 |

†*Unadjusted p-values. Values indicated as "0.000" are smaller than 0.0001. Orange highlighted rows compare post-IRA proportion to pre-IRA proportion*


© Vital Transformation, LLC 2026

#### Table A7 - Contrasts of predicted proportion of hematology secondary indication studies by modality

| Contrast | | Difference in Predicted Proportions | Std Err | Z-stat | | p-value† |
|---|---|---|---|---|---|---|
| Pre-IRA Large vs Pre-IRA Small | | -0.074 | 0.042 | -1.76 | | 0.078 |
| Post-IRA Small vs Pre-IRA Small | | 0.077 | 0.033 | 2.32 | | 0.020 |
| Post-IRA Large vs Pre-IRA Small | | -0.097 | 0.037 | -2.64 | | 0.008 |
| Post-IRA Small vs Pre-IRA Large | | 0.151 | 0.043 | 3.55 | | 0.000 |
| Post-IRA Large vs Pre-IRA Large | | -0.023 | 0.045 | -0.51 | | 0.609 |
| Post-IRA Large vs Post-IRA Small | | -0.174 | 0.037 | -4.67 | | 0.000 |

#### Table A8 - Contrasts of predicted proportion of hematology secondary indication studies by rare disease status

| Contrast | Difference in Predicted Proportions | Std Err | Z-stat | p-value† |
|---|---|---|---|---|
| Pre-IRA Orphan vs Pre-IRA Non-Orphan | 0.013 | 0.011 | 1.22 | 0.222 |
| Post-IRA Non-Orphan vs Pre-IRA Non-Orphan | 0.008 | 0.009 | 0.9 | 0.368 |
| Post-IRA Orphan vs Pre-IRA Non-Orphan | 0.044 | 0.014 | 3.19 | 0.001 |
| Post-IRA Non-Orphan vs Pre-IRA Orphan | -0.005 | 0.011 | -0.43 | 0.665 |
| Post-IRA Orphan vs Pre-IRA Orphan | 0.031 | 0.016 | 1.95 | 0.051 |
| Post-IRA Orphan vs Post-IRA Non-Orphan | 0.036 | 0.014 | 2.62 | 0.009 |

*†Unadjusted p-values. Values indicated as "0.000" are smaller than 0.0001. Orange highlighted rows compare post-IRA proportion to pre-IRA proportion*


© Vital Transformation, LLC 2026
